# Investigating the impact of London’s Ultra Low Emission Zone (ULEZ) on children’s health: cohort description and baseline data from the Children’s Health in London and Luton (CHILL) prospective parallel cohort study

**DOI:** 10.1101/2024.06.05.24308482

**Authors:** Helen E. Wood, Harpal Kalsi, Louise Cross, Rosamund E. Dove, James Scales, Ivelina Tsocheva, Jasmine Chavda, Grainne Colligan, Esther Lie, Kristian Petrovic, Florian Tomini, Veronica Tofolutti, Bill Day, Amanda Keighley, Cheryll Critchlow, Sean Beevers, Monica Fletcher, W. James Gauderman, Jonathan Grigg, Borislava Mihaylova, Chris Newby, Esther van Sluijs, Frank Kelly, Aziz Sheikh, Gurch Randhawa, Ian S. Mudway, Chris J. Griffiths

## Abstract

Traffic-related air pollution (TRAP) poses significant health risks particularly for children, with adverse effects that may impact health in later life. Low emission zones are a public health policy designed to reduce TRAP in urban areas.

The CHILL (Children’s Health in London and Luton) Study will evaluate the impact of London’s Ultra Low Emission Zone (ULEZ) on children’s health, using a prospective two-arm parallel longitudinal cohort design. CHILL will examine associations between air pollution metrics and lung function growth, plus secondary outcomes. Here we describe the characteristics of the CHILL cohort at baseline, prior to the introduction of the ULEZ.

We recruited 3414 children aged 6-9 years attending 84 schools (London – intervention site: 1664 children, 44 schools; Luton – comparator site: 1750 children, 40 schools). Baseline health assessments were conducted in 2018-2019 (before the introduction of the ULEZ in London). 97.0% of recruited children were assessed (London 96.5%, Luton 97.4%), with the primary outcome measure of post-bronchodilator forced expiratory volume in one second being successfully measured in 76.7% (London 76.9%, Luton 76.5%). 92.1% returned a completed parental questionnaire (London 89.3%, Luton 94.7%), including data for analysis of the secondary outcomes. Demographic characteristics and outcomes were similar across the two sites.

We established well-matched cohorts of school children, in our intervention (London) and comparator (Luton) sites. Data on primary and secondary outcomes have been successfully collected, which, combined with detailed air quality metrics, provides a robust platform for evaluating the impact of London’s ULEZ on children’s health and development.

ClinicalTrials.gov NCT04695093 (05/01/2021)

## INTRODUCTION

Air pollution contributes to over 7 million deaths per year across the globe (1) and poses the single largest environmental risk to health (2). In Europe, nearly 800,000 excess deaths a year are caused, in part, by poor air quality in densely populated urban areas (3). In the UK, the majority of under-fives, young adults and poorer households live in areas with the highest concentrations of traffic-related air pollution (TRAP) (4), resulting in a disproportionate health burden falling on some of the most disadvantaged and vulnerable members of society (5) and disproportionate numbers of poor and vulnerable people being affected (6). Children are particularly sensitive to the harmful effects of air pollution (7) and concern has increased over recent years that long-term exposures can cause developmental delays with long-term consequences on children’s health across the life course (8).

Stringent clean air targets recently set by the World Health Organisation urgently need to be implemented to reduce the exposure of children to air pollution (9). Clean air zones (CAZs) are areas where special measures have been put in place to improve air quality and may be an essential tool in meeting proposed air quality targets. The ultra low emission zone (ULEZ) is a clean air zone that restricts and penalises the entry of high-polluting vehicles (based on European class emission standards) into central London. It is hoped that the introduction of the ULEZ will keep the most polluting vehicles out of the area, improve air quality and encourage positive changes in modes of transport.

The Children’s Health in London and Luton (CHILL) Study uses a prospective parallel cohort design to investigate whether the introduction of London’s ULEZ has delivered a beneficial impact on the health of children living in central London. The cohort is comprised of two closely matched groups of primary school children residing in central London (intervention area), and Luton (comparison area). We anticipate that data from the CHILL Study will contribute epidemiological evidence to assess the impact of air pollution on childhood development and the extent to which environmental regulatory policies, such as the ULEZ, may improve health (10), with implications for local, national and international policy.

Primary school-aged children were recruited to the CHILL Study during 2018-2019, supported by a school outreach programme, with cohort recruitment in London being completed immediately prior to the implementation of the ULEZ on 8^th^ April 2019 (recruitment in Luton was completed shortly thereafter). We present here baseline demographic, health, and air quality exposure data for the CHILL cohort, including lung function, respiratory health, and quality of life.

## METHODS

### Study design

The CHILL Study design and methods have been reported previously (11, 12). Briefly, CHILL is a prospective parallel longitudinal cohort study performed over a 5-year period at two locations: the intervention site impacted by the ULEZ road traffic management scheme (Central London) and the comparator site (Luton). The study design is summarised in Fig. 1. The duration of data collection now extends to include a 5^th^ year as a result of the COVID-19 pandemic, hence this diagram updates that previously published (13). The configurations of the ULEZ at its commencement in central London in 2019, and when it was extended in 2021 to encompass inner London, are shown in Fig. 2.

**Fig. 1.**
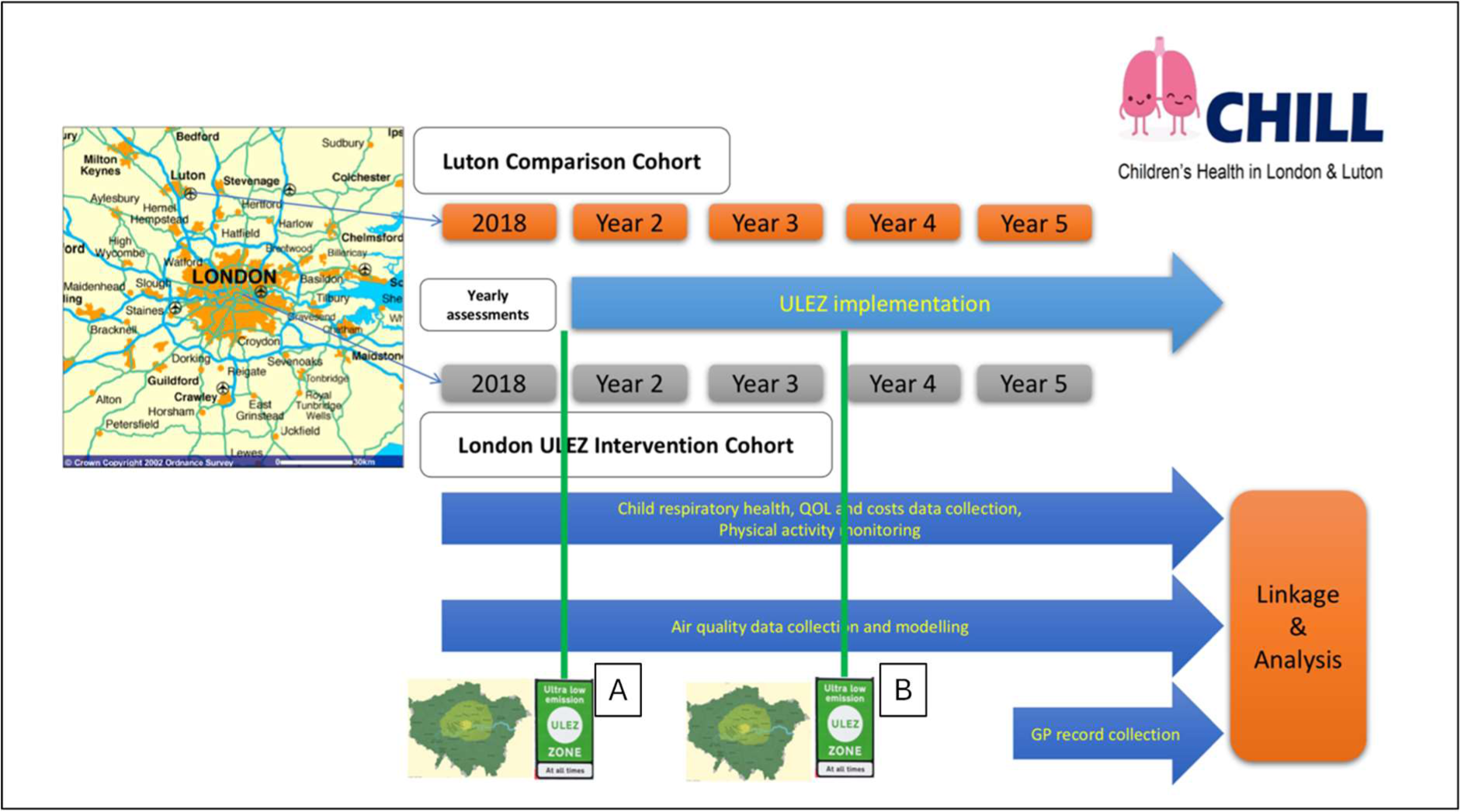
Study scheme diagram. The figure illustrates the CHILL study design with green lines indicating the implementation of the ULEZ and its subsequent expansion: (A) The introduction of the original ULEZ to central London only, in April 2019; (B) the extension of the ULEZ to the boundaries of the North (A406) and South Circular Roads (A205) in October 2021. Data collection from participants was carried out annually, beginning in 2018/2019, prior to the introduction of the original ULEZ, and ended prior to the second expansion of the scheme to cover all London boroughs in August 2023. Data collection from GP records will be carried out in the final year of the study

**Fig. 2.**
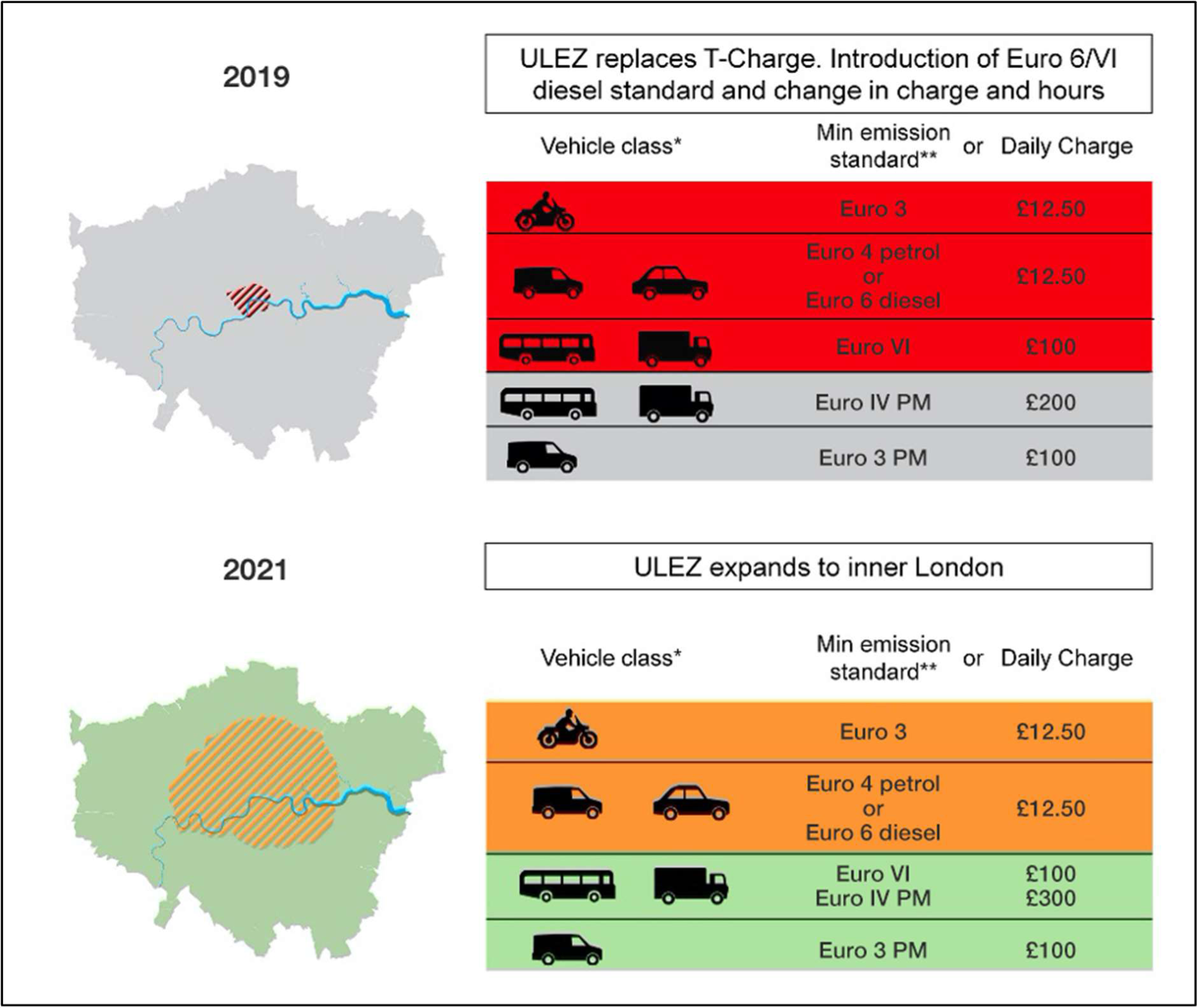
ULEZ configuration during phase 1 (2019) and the subsequent extension in 2021. Panels illustrate the size of the area covered by the ULEZ from 8^th^ April 2019 and 25^th^ October 2021, with the minimum Euro emission standards for different vehicle classes illustrated, with the daily charges for non-compliant vehicles

### Study population

All primary schools located within the central London ULEZ area, or with a catchment area including the ULEZ, and all those within the borough of Luton were invited to participate in the study. In Luton, recruitment was extended to include primary schools in the contiguous neighbouring town of Dunstable to enable sufficient participant numbers. 44 primary schools in central London (67% of those invited) and 40 in Luton/Dunstable (32 in Luton and 8 in Dunstable, 71% of those invited; henceforth referred to as ‘Luton’, for simplicity) agreed to participate. Depending on the size of the school, all children in school years 2, 3 and 4 (those aged 6-9 years) were given a study pack to take home (including study information sheets, a consent form and a questionnaire to be completed by their parent/carer). In some larger schools with more than three classes per year group, packs were only sent to one or two year groups; this was more often the case in Luton, where schools tended to be larger than in London. Children who returned a signed consent form to school before the scheduled health assessment visit were recruited to the study. Inclusion/exclusion criteria are published elsewhere (11). A target of 3200 participants (1600 each in London and Luton) and 40 schools from each site was required to meet sample size requirements (11).

Recruitment commenced in June 2018 to permit collection of a year of pre-ULEZ baseline data. In London, recruitment and baseline health assessments were completed on April 6^th^ 2019, two days prior to the introduction of the ULEZ. In Luton, recruitment and baseline health assessments continued until 26^th^ April 2019, to achieve the target sample number of participants. In London, a total of 4535 children were invited to participate in the CHILL Study, of which 1664 returned a signed consent form. In Luton, 4884 children were invited to take part and 1750 of these returned a signed consent form.

### Data collection

Data were collected directly from children at health assessments visits to their school and indirectly from the questionnaire completed by their parent/carer and returned to school. The primary outcome variable for this study was forced expiratory volume in 1 second (FEV_1_) as measured by spirometry, secondary outcome variables are summarised in Table 1.

**Table 1.** Data collected directly from children during health assessments, and indirectly from parent/carer-completed questionnaires.

| Health assessment | Questionnaire |
| --- | --- |
| Height, seated height, weight | Ethnicity |
| Lung function (FEV<sub>1</sub><sup>a</sup>, FVC<sup>b</sup> and other spirometric variables) | Home address and address history |
| Physical activity (Accelerometry) and GPS tracking<sup>c</sup> | Parent-reported mode of travel to school |
| Self-reported asthma, inhaler use | Exposure to second-hand smoke |
| Self-reported mode of travel to school | Respiratory and allergic symptoms (ISAAC<sup>d</sup>) |
|  | Health-related quality of life (CHU9D<sup>e</sup>) |
|  | School absence due to ill health, parental work absence |
|  | Non-NHS health-related costs |
|  | GP details, NHS number |

<sup>a</sup> forced expiratory volume in one second, <sup>b</sup> forced vital capacity, <sup>c</sup> subset of children; all those taking part in the health assessment were offered a physical activity monitor, while all those in Year 3 at baseline were offered a Global Positioning System (GPS) monitor (except in circumstances where insufficient monitors were available), <sup>d</sup> ISAAC: International Study of Asthma and Allergy in Children Questionnaire (14), <sup>e</sup> CHU9D: Child Health Utility 9D Questionnaire (15)

#### Index of Multiple Deprivation (IMD)

Children’s home addresses at the time of the baseline heath assessment were collected via parent/carer questionnaires. Home postcodes were mapped to IMD decile using the English Indices of Deprivation 2019 (16).

#### Lung function

After measurement of height and weight, lung function was measured using a Vitalograph 6000 Alpha Touch Spirometer (Vitalograph, Buckingham, UK), before and 15 minutes after administration of a bronchodilator (salbutamol), according to American Thoracic Society and European Respiratory Society (ATS/ERS) 2005 guidelines (17, 18). Spirometers were calibrated with a three-litre precision syringe, and biological control assessments were performed, prior to each assessment session. Children inhaled four puffs (100mcg/puff) of salbutamol *via* a large volume spacer (Volumatic, Allen and Hanburys Ltd, UK), administered by a study team member. A new or disinfected spacer was used for each child.

Quality assurance of data was performed according to guidelines (18). In summary, spirometry was deemed successful if the participant produced three acceptable measures of forced vital capacity (FVC) and FEV_1_ within 150ml, free from artefacts at all stages, with a plateau in the flow/time loop profile. Measures could be accepted if there were fewer than three attempts within 150ml, at the discretion of the Senior Respiratory Physiologist (HK) if there was suitable evidence that all manoeuvres had been performed maximally and without artefact. Spirometry variables to be included in the analysis are: FEV_1_, FVC, FEV_1_/FVC ratio, peak expiratory flow (PEF), and (maximal-mid expiratory flow between 75 and 25% of the forced vital capacity) MMEF_75/25_.

#### Physical activity and GPS tracking

Physical activity was measured using ActiGraph triaxial accelerometers (ActiGraph wGT3X-BT, Pensacola, FL USA), which were given to the children along with verbal and written instructions on their use. Children were asked to wear the accelerometer during waking hours for seven days, on their right hip using an elastic belt, after which they were to return it to school for collection by the study team. Valid daily wear time at baseline was set at five consecutive days of 480 minutes between 6am and 11pm. Accelerometer data were downloaded and averaged over five-second epochs using the ActiLife software programme for initial data checking. Data files with hourly-level data were subsequently processed in Stata (Version 13, StataCorp. College Station, TX, USA) to remove periods of 60 minutes or more of continuous zero acceleration.

Children in Year 3 (aged 7-8 years) at baseline were also asked to wear a GPS monitor (QStarz BT-Q1000XT, Taipei, Taiwan) for the same seven-day period, to assess the routes used to travel to and from school and, combined with accelerometry, to provide an objective measure of travel mode to school. Follow-up assessments with GPS monitors were only conducted in this sub-cohort.

#### Self-reported inhaler use and travel

At the start of the health assessment, children were asked whether they have asthma and whether they used any inhalers. If answering yes, they were asked which type of inhaler(s) they used and whether they had used any that day, prior to the health assessment. If a child indicated they had already used a bronchodilator that day (salbutamol or ‘blue’ inhaler), further bronchodilator was not given during the health assessment and spirometry was only measured once, that measurement being assumed to be ‘post-bronchodilator’(19). Children were additionally asked about how they travelled to school that day and how they would usually travel.

#### Parent/carer-completed questionnaire

Prior to a school health assessment visit, children were given a study pack to take home, including a questionnaire which parents/carers were asked to complete and return to school. If not returned prior to the health assessment visit, completed questionnaires were collected by the study team at follow-up visits to the school and/or a second copy was provided if the original had been mislaid. Questions were included on the topics indicated in Table 1.

Data from returned questionnaires was entered by researchers into the Research Electronic Data Capture (REDCap) application, a secure site for managing clinical research data, and was overseen and quality controlled by the Pragmatic Clinical Trials Unit (PCTU) at Queen Mary University of London and the CHILL Study project managers. Questionnaire responses were entered into the database as recorded on the hard copy, regardless of inconsistencies. Where questions were left unanswered, a “no answer” field was selected to account for missing data.

### Air quality exposures

Luton/Dunstable was chosen as the comparator site to London because it met the six target criteria:

1. be geographically distant and therefore free from contamination from the ULEZ.
2. have a similar population demographic to London.
3. have no plans to enact any form of low emission zone.
4. have a similar mixture of local pollutant sources to Central London.
5. have continuous air quality (AQ) monitoring sites, with freely available long-term trend data.
6. have annual nitrogen dioxide (NO_2_) concentrations at the urban background of >40ug/m^3^, with clear exceedances at roadside locations.

Luton is a large urban town of 214,600 population, 34 miles from London. Its population has an ethnic mix of 54.6% White, 30.0% Asian, 9.8% Black, 4.1% Mixed Race, 1.5% other ethnic groups (20). Luton’s AQ is influenced by several factors: the presence of major industry (including a motor industry), the transecting M1 motorway, and a rapidly expanding international airport, all bringing significant traffic flows into/through the town. Luton’s AQ strategy at the start of the CHILL Study is summarised in its 2018 report (21). It has three designated AQ Management Areas. Mean annual NO_2_ values have remained largely unchanged over the five years prior to the start of the study. Luton Council’s planned interventions for AQ improvement included a busway, car sharing, public information and advice systems, a scheme promoting cycling by commuters to railway stations, and the provision of a limited number of electric vehicle charging points (22). It had no plans for a Clean Air Zone. This AQ strategy, the limited control Luton has over traffic emissions from the M1, and its expanding airport mean large improvements in AQ were unlikely in the next five years.

## RESULTS

### Cohort recruitment and baseline data collection

We achieved a cohort size slightly over the recruitment target, as shown in Fig. 3, which provides a breakdown of schools, participants, returned parent/carer questionnaires and numbers for participants completing lung function assessments at baseline across both sites.

**Fig. 3.**
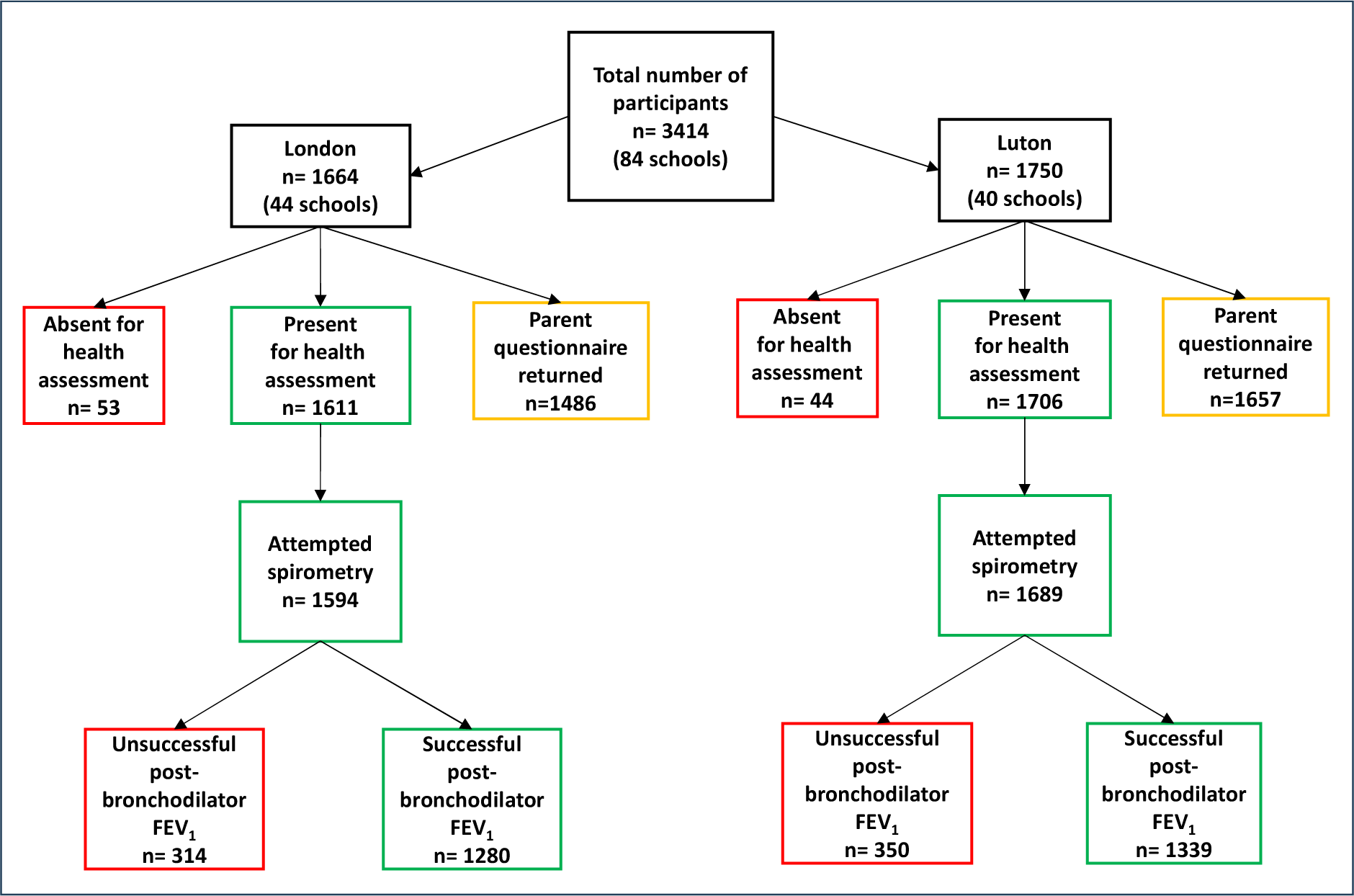
Cohort recruitment and completeness of baseline data collection. FEV_1_, forced expiratory volume in 1 second.

### Demographics and lung function

Summary demographic data, comparing the London and Luton cohorts are shown in Fig. 4. There was some disparity between the two sites in the distribution of participants by school year, such that the average age was slightly higher in London than Luton (7.95 ± 0.89 versus 7.78 ± 0.80 years, mean ± SD). There were more girls than boys recruited in London (55.2 vs. 44.8%), while in Luton the proportions of girls and boys were similar (49.1 vs. 50.9%). Across both sites, the proportions of children from ethnic minorities were similar, but the distribution of ethnic groups was different, with a higher proportion of Asian/Asian British children recruited in Luton (38.4 vs. 20.7% in London) and a higher proportion of Black/Black British children recruited in London (18.9 vs. 7.5% in Luton).

**Fig. 4.**
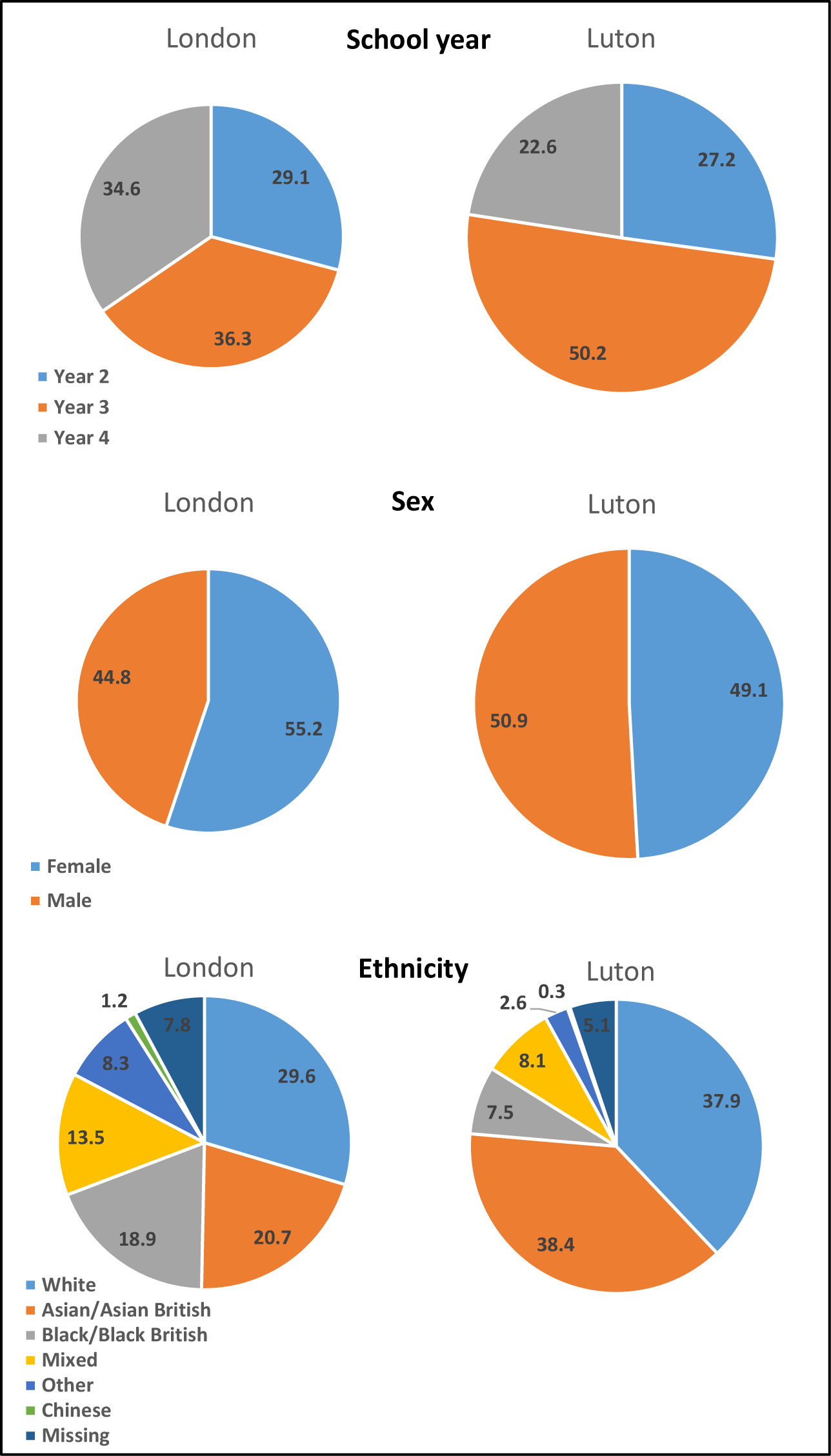
Demographics of the recruited cohort in each site, by school year group, sex, and ethnicity.

Summary data from the baseline health assessment are shown in Table 2 for the whole cohort and individually for London and Luton. A small proportion (around 3%) of the cohort were either absent for the baseline health assessment visit or declined to participate on the day, with height and weight being measured for 1606 participants in London and 1705 in Luton. Despite the differences in distribution of age, sex, and ethnic groups between the two sites, average height, weight, and BMI were similar in London and Luton.

**Table 2.** Baseline health assessment data: data completeness and average anthropometric measurements for the whole cohort and by site.

| Variable | All<br>N (%)<sup>a</sup> | London | Luton |
| --- | --- | --- | --- |
| Present for health assessment, n (%)<sup>a</sup> | 3317 (97.2) | 1611 (96.8) | 1706 (97.5) |
| Absent for health assessment, n (%) | 97 (2.8) | 53 (3.2) | 44 (2.5) |
| Declined to take part in health assessment, n (%) | 6 (0.2) | 5 (0.3) | 1 (0.1) |
| Height and weight measured, n (%) | 3311 (97.0) | 1606 (96.5) | 1705 (97.4) |
| Height (cm), Mean +/- SD | 128.4 +/- 7.7 | 129.3 +/- 8.1 | 127.6 +/- 7.4 |
| Height (cm), Min/Max | 107.0 / 156.6 | 107.1 / 156.6 | 107.0 / 155.5 |
| Weight (kg), Mean +/- SD | 28.5 +/- 7.0 | 29.0 +/- 7.3 | 28.0 +/- 6.8 |
| Weight (kg), Min/Max | 14.8 / 79.8 | 15.2 / 79.8 | 14.8 / 63.2 |
| BMI (kg/m<sup>2</sup>), Mean +/- SD | 17.1 +/- 2.8 | 17.2 +/- 2.9 | 17.0 +/- 2.8 |
| BMI (kg/m<sup>2</sup>), Min/Max | 8.3 / 34.2 | 12.0 / 34.2 | 8.3 / 31.6 |
| Attempted spirometry, n (%) | 3283 (96.2) | 1594 (95.8) | 1689 (96.5) |
| Successful post-bronchodilator FEV<sub>1</sub>, n (%) | 2619 (76.7) | 1280 (76.9) | 1339 (76.5) |
| Successful post-bronchodilator FVC, n (%) | 2577 (75.5) | 1255 (75.4) | 1322 (75.5) |
| Successful pre- and post- FEV<sub>1</sub>, n (%) | 2452 (71.8) | 1190 (71.5) | 1262 (72.1) |
| Successful pre- and post- FVC, n (%) | 2411 (70.6) | 1167 (70.1) | 1244 (71.1) |

<sup>a</sup> percent of recruited participants; BMI, body mass index; FEV<sub>1</sub>, forced expiratory volume in 1 second; FVC, forced vital capacity; SD, standard deviation

Spirometry was attempted by 96.2% of the cohort (3283/3414), with 76.7% providing successful measurement of the primary outcome variable (2619/3414, post bronchodilator FEV_1_). Successful measurement of FEV_1_ both pre- and post-bronchodilator, was achieved for 71.8% of the cohort (2452/3414), permitting calculation of reversibility. Unsuccessful spirometry was due to several reasons: some children were unable to perform the FVC manoeuvre, some declined to receive salbutamol, and some had already taken a bronchodilator inhaler on the day and their spirometry was deemed as ‘post-bronchodilator’ (these children provided usable post-bronchodilator measures, but not reversibility).

Post-bronchodilator spirometry variables are shown in Table 3, for the whole cohort and each site. Variables are shown as absolute values and as percent of predicted, based on age, height, weight and assigned ethnic population, as categorised by the Global Lung Initiative (23) (Caucasian, Afro-Caribbean, Northeast Asian, Southeast Asian and Mixed Race/Other). Categorisation was established by asking participants where they were born and where their parents were born. Averaged across all children in each site, baseline lung function was similar between London and Luton, both in absolute terms and as a percent of predicted values. FEV_1_, the main outcome measure of the study, was almost identical between the two sites (1.57 ± 0.30 vs. 1.57 ± 0.29 L, in London and Luton, respectively), as was FVC (1.79 ± 0.35 vs. 1.81 ± 0.34 L). The only notable exception was the change in FEV_1_ from pre- to post-bronchodilator measurement, which was greater in London (5.70 +/- 9.37 vs. 5.03 +/- 8.72 % in Luton), indicating higher average reversibility, and potentially inferring a greater degree of undiagnosed asthma in London.

**Table 3.**
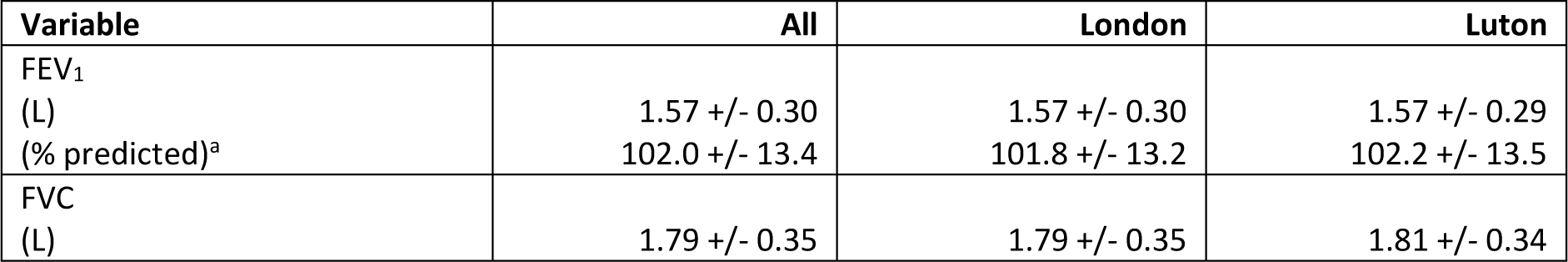
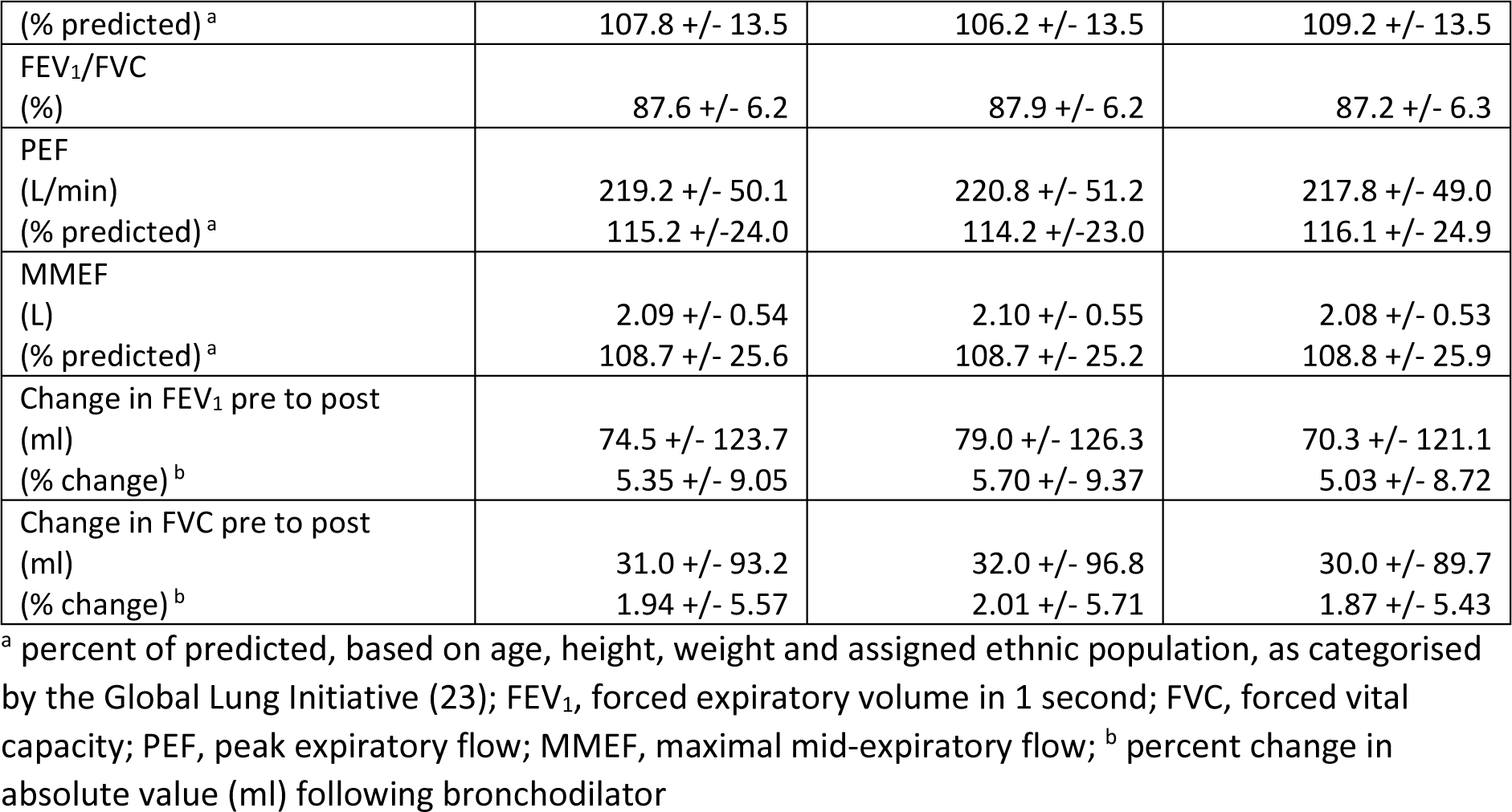
Baseline post-bronchodilator spirometric measurements for the whole cohort and by site, data are mean +/- standard deviation.

Baseline postcode was available for 3271 participants (95.8%), although 149 postcodes failed to match with the IMD database, leaving 3122 participants (91.4%) with IMD assigned (London, n=1475; Luton, n=1647). IMD decile was higher on average in Luton, and there was also a greater amount of variability (London, 3.58 +/- 1.69; Luton, 4.71 +/- 1.87; mean +/- SD). Similar (very small) proportions of participants lived in areas ranked in the highest and lowest deciles for IMD, i.e., least and most deprived, in London and Luton (least deprived, 0.6 and 0.5%, respectively; most deprived 1.5 and 1.3%, respectively). However, higher proportions lived in the 2^nd^, 3^rd^ and 4^th^ most deprived deciles in Luton compared with London (see Fig. 5).

**Fig. 5.**
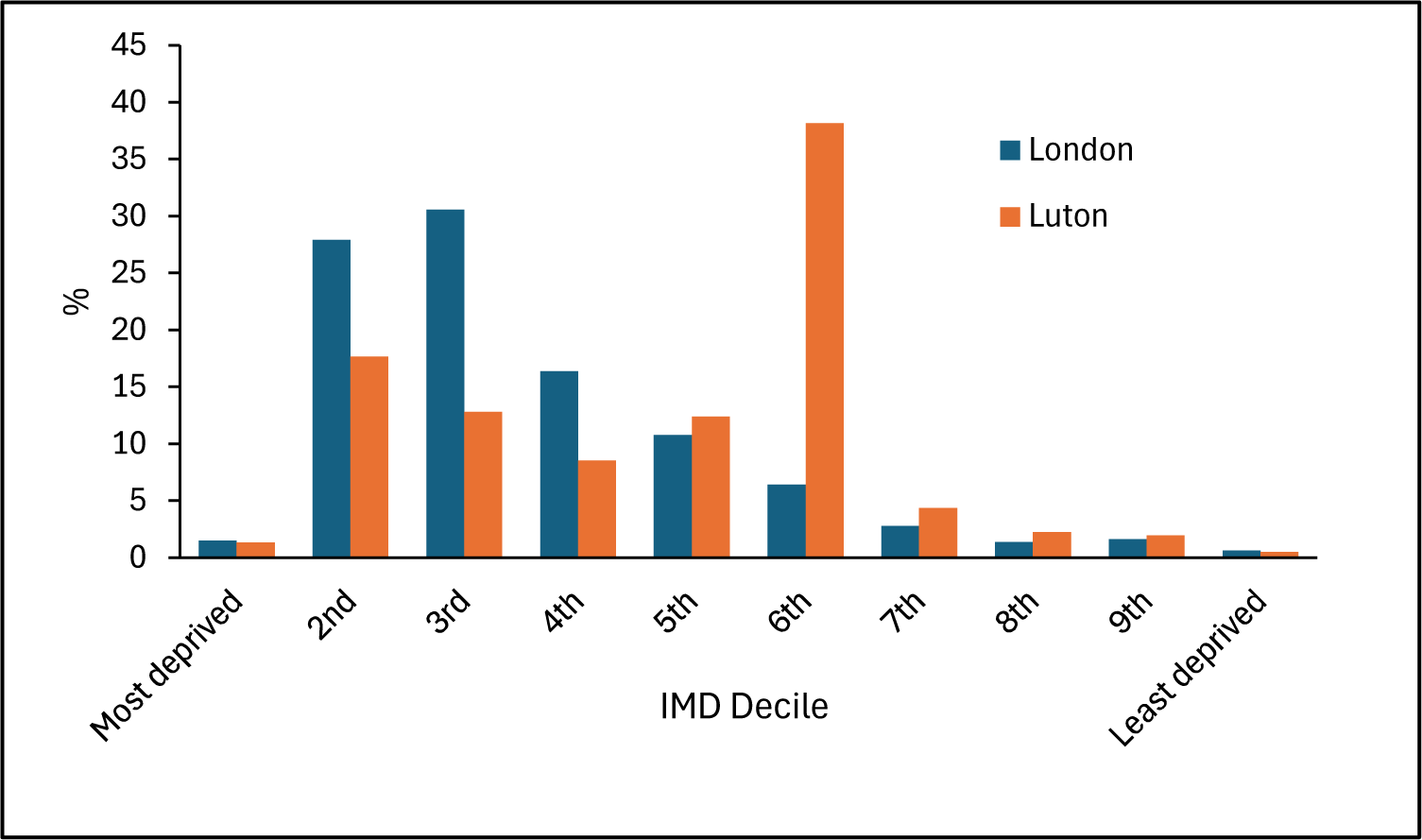
Distribution of cohort, by site, across deciles of deprivation (IMD 2019), data are percentage of participants with IMD assigned.

### Self-reported inhaler use and travel

Data on self-reported use of inhalers and travel to school, from the 3311 children who participated in the baseline health assessment visits, are shown in Table 4. Asthma was self-reported by 10.9% of children (361/3311), with 13.4% (444/3311) reporting inhaler use. The most used type was the ‘blue’ (salbutamol) inhaler, reported by 93.2% of inhaler-users (414/444). Self-reported asthma and inhaler use were slightly higher in Luton as compared with London (12.6 vs. 9.1% and 14.5 vs. 12.3%, respectively) (see Table 5).

**Table 4.** Self-reported asthma and inhaler use, and method of travel to school, by children participating in the health assessment visits.

| Variable | All | London | Luton |
| --- | --- | --- | --- |
| Participated in health assessment (N, % of cohort) <sup>a</sup> | 3311 (97.0) | 1606 (96.5) | 1705 (97.4) |
| Self-reported asthma | 361 (10.9) | 146 (9.1) | 215 (12.6) |
| Self-reported inhaler use | 444 (13.4) | 197 (12.3) | 247 (14.5) |
| Self-reported blue inhaler use<sup>b</sup> | 414 (93.2) | 183 (92.9) | 231 (93.5) |
| Self-reported brown inhaler use<sup>b</sup> | 128 (28.8) | 61 (31.0) | 67 (27.1) |
| Self-reported other inhaler use<sup>b</sup> | 39 (8.8) | 22 (11.2) | 17 (6.9) |
| Self-reported method of travel to school on day of health assessment<sup>c</sup>: |  |  |  |
| Walk | 2012 (60.8) | 1069 (66.6) | 943 (55.3) |
| Scoot | 144 (4.3) | 114 (7.1) | 30 (1.8) |
| Bike | 62 (1.9) | 43 (2.7) | 19 (1.1) |
| Private Car | 976 (29.5) | 200 (12.5) | 776 (45.5) |
| Taxi | 22 (0.7) | 7 (0.4) | 15 (0.9) |
| Bus | 231 (7.0) | 207 (12.9) | 24 (1.4) |
| Train/Tube | 53 (1.6) | 53 (3.3) | 0 (0.0) |
| Self-reported usual method of travel to school<sup>c</sup>: |  |  |  |
| Walk | 2526 (76.3) | 1256 (78.2) | 1270 (74.5) |
| Scoot | 404 (12.2) | 245 (15.3) | 159 (9.3) |
| Bike | 233 (7.0) | 117 (7.3) | 116 (6.8) |
| Private Car | 1317 (39.8) | 323 (20.1) | 994 (58.3) |
| Taxi | 53 (1.6) | 18 (1.1) | 35 (2.1) |
| Bus | 327 (9.9) | 284 (17.7) | 43 (2.5) |
| Train/Tube | 71 (2.1) | 71 (4.4) | 0 (0.0) |

<sup>a</sup> percentages are of those who participated in the health assessment, unless otherwise specified; <sup>b</sup> percent of those who reported inhaler use; <sup>c</sup> percent totals add to more than 100 because children could give more than one answer

**Table 5.** Prevalence of respiratory and allergic symptoms, as well as exposure to cigarette smoke at home, from parent/carer questionnaire, reported as percentage of those who returned a completed questionnaire, unless otherwise specified.

| Variable | All | London | Luton |
| --- | --- | --- | --- |
| Returned parent questionnaire (N, % of cohort) | 3143 (92.1) | 1486 (89.3) | 1657 (94.7) |
| Reported cigarette smoker in the household | 700 (22.3) | 264 (17.8) | 436 (26.3) |
| Reported their child had wheezing in last 12 months | 357 (11.4) | 162 (10.9) | 195 (11.8) |
| Reported their child had severe wheeze | 145 (4.6) | 65 (4.4) | 80 (4.8) |
| Reported their child ever had asthma | 377 (12.0) | 150 (10.1) | 227 (13.7) |
| Reported their child experienced exercise-induced wheeze in last 12 months | 239 (7.6) | 107 (7.2) | 132 (8.0) |
| Reported their child had dry cough at night in last 12 months, not associated with a cold | 626 (19.9) | 303 (20.4) | 323 (19.5) |
| Reported their child had problem with sneezing or runny/blocked nose in last 12 months, not associated with a cold | 680 (21.6) | 368 (24.8) | 312 (18.8) |
| Reported their child also had itchy-watery eyes in the last 12 months | 359 (11.4) | 196 (13.2) | 163 (9.8) |
| Reported their child ever had hayfever | 888 (28.3) | 435 (29.3) | 453 (27.3) |
| Reported their child had eczema in last 12 months | 423 (13.5) | 215 (14.5) | 208 (12.6) |
| Reported their child ever had eczema | 868 (27.6) | 426 (28.7) | 442 (26.7) |
| Reported their child had current wheeze, rhinitis and eczema | 69 (2.2) | 40 (2.7) | 29 (1.8) |

Across both sites, walking was the most common method of travel to school, both on the day of the health assessment and ‘usually’, reflecting the proximity of primary schools to participants’ home addresses in many cases. More participants in London reported using active travel methods (walking, scooting, cycling) and public transport (train, tube), whereas participants in Luton were more likely to travel to school by private car. Note that for these questions, children could give more than one answer, so totals do not add up to 100% (see Table 4).

### Respiratory and allergy symptoms

The Baseline parent/carer questionnaire was returned by 92% of the cohort overall (3143/3414), with a higher return rate in Luton, as compared with London (94.7% vs. 89.3%, 1657/1750 vs. 1486/1664). Data for the prevalence of respiratory and allergy symptoms (collected using the ISAAC questionnaire, (14)) are shown in Table 5. A higher proportion of respondents reported a cigarette smoker in the household in Luton than London (26.3% vs. 17.8%, 436/1657 vs. 264/1486). In Luton, there were also slightly higher proportions who reported their child wheezing in the last 12 months (11.8 vs. 10.9% in London, 195/1657 vs. 162/1486) and having ever had asthma (13.7 vs. 10.1% in London, 227/1657 vs. 150/1486). On the other hand, rhinitis symptoms (sneezing/runny/blocked nose: 24.8 vs. 18.8% in Luton, 368/1486 vs. 312/1657; itchy-watery eyes: 13.2 vs. 9.8% in Luton, 196/1486 vs. 163/1657) and having ever had hay fever (29.3 vs. 27.3% in Luton, 435/1486 vs. 453/1657) were more common in London. The incidence of eczema was similar across sites.

### Quality of life

Health-related quality of life (QoL) data at Baseline are shown in Table 6, based on the Child Health Utility 9D (CHU9D) Questionnaire (15), included in the parent/carer questionnaire. Children scored relatively high in each of the domains, except for feeling tired (12.2% of the children in London and 11.8% in Luton felt a bit, quite, or very tired), feeling annoyed (5.7% in London and 5.6% in Luton felt a bit, quite, or very annoyed), school work/homework (6.7% in London and 6.2% in Luton had some, many problems or could not do school/homework) and sleeping (5.5% in London and 4.2% in Luton had at least some difficulties with sleeping).

**Table 6.**
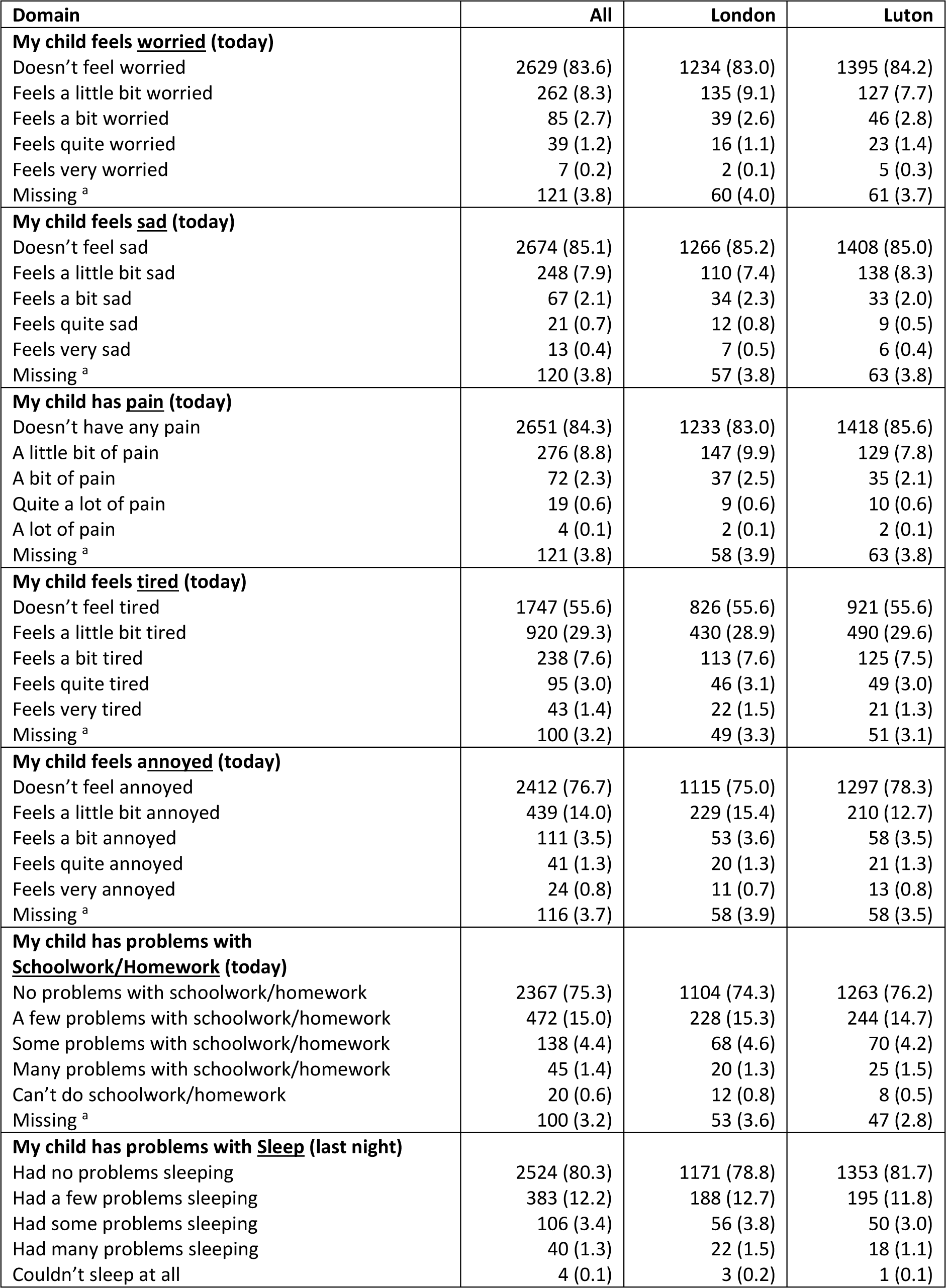
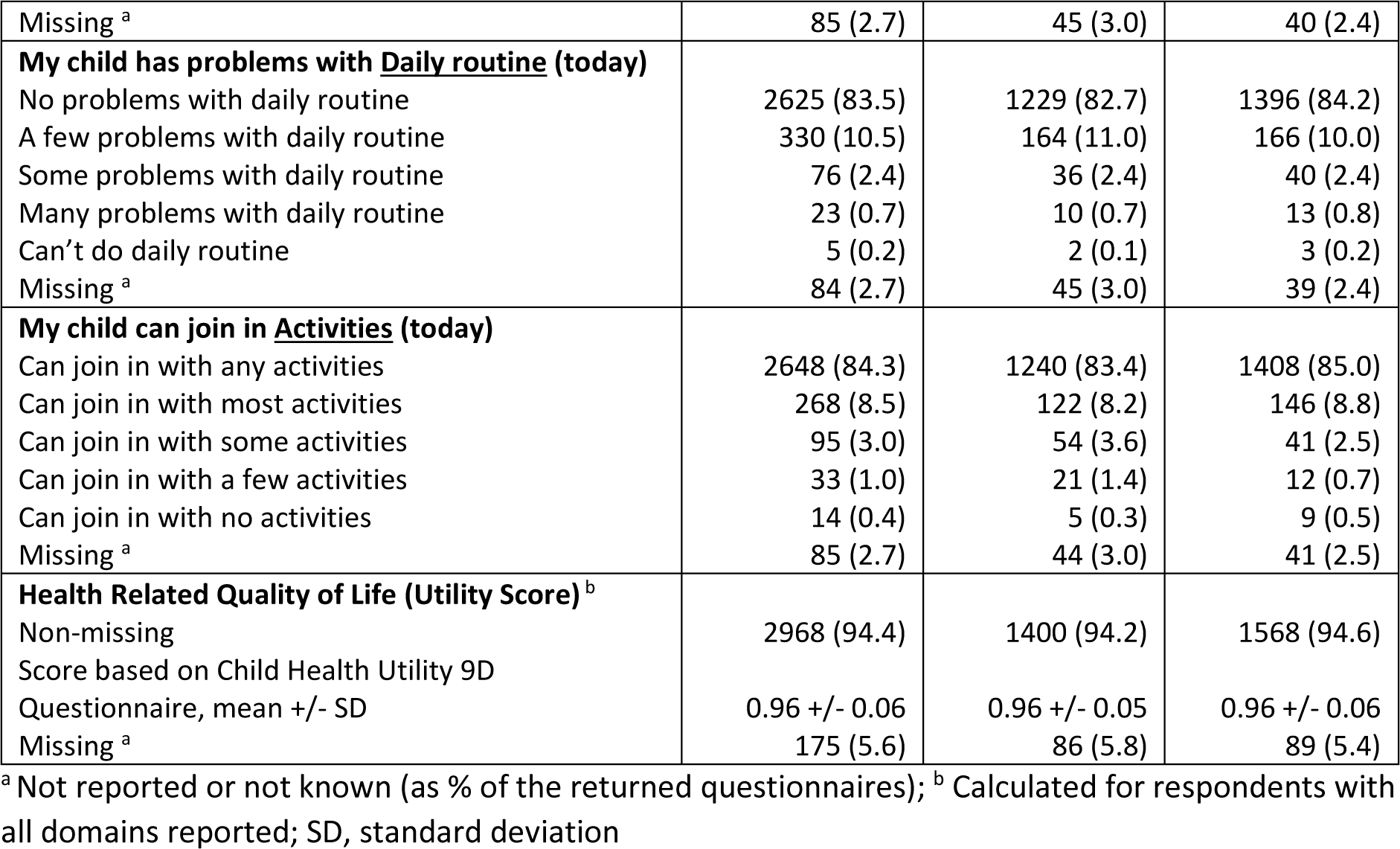
Health Related Quality of Life of primary-aged school children in London and Luton based on Child Health Utility 9D Questionnaire, data are N (% of those who returned a questionnaire)

| Domain | All | London | Luton |
| --- | --- | --- | --- |
| **My child feels <u>worried</u> (today)** |  |  |  |
| Doesn't feel worried | 2629 (83.6) | 1234 (83.0) | 1395 (84.2) |
| Feels a little bit worried | 262 (8.3) | 135 (9.1) | 127 (7.7) |
| Feels a bit worried | 85 (2.7) | 39 (2.6) | 46 (2.8) |
| Feels quite worried | 39 (1.2) | 16 (1.1) | 23 (1.4) |
| Feels very worried | 7 (0.2) | 2 (0.1) | 5 (0.3) |
| Missing <sup>a</sup> | 121 (3.8) | 60 (4.0) | 61 (3.7) |
| **My child feels <u>sad</u> (today)** |  |  |  |
| Doesn't feel sad | 2674 (85.1) | 1266 (85.2) | 1408 (85.0) |
| Feels a little bit sad | 248 (7.9) | 110 (7.4) | 138 (8.3) |
| Feels a bit sad | 67 (2.1) | 34 (2.3) | 33 (2.0) |
| Feels quite sad | 21 (0.7) | 12 (0.8) | 9 (0.5) |
| Feels very sad | 13 (0.4) | 7 (0.5) | 6 (0.4) |
| Missing <sup>a</sup> | 120 (3.8) | 57 (3.8) | 63 (3.8) |
| **My child has <u>pain</u> (today)** |  |  |  |
| Doesn't have any pain | 2651 (84.3) | 1233 (83.0) | 1418 (85.6) |
| A little bit of pain | 276 (8.8) | 147 (9.9) | 129 (7.8) |
| A bit of pain | 72 (2.3) | 37 (2.5) | 35 (2.1) |
| Quite a lot of pain | 19 (0.6) | 9 (0.6) | 10 (0.6) |
| A lot of pain | 4 (0.1) | 2 (0.1) | 2 (0.1) |
| Missing <sup>a</sup> | 121 (3.8) | 58 (3.9) | 63 (3.8) |
| **My child feels <u>tired</u> (today)** |  |  |  |
| Doesn't feel tired | 1747 (55.6) | 826 (55.6) | 921 (55.6) |
| Feels a little bit tired | 920 (29.3) | 430 (28.9) | 490 (29.6) |
| Feels a bit tired | 238 (7.6) | 113 (7.6) | 125 (7.5) |
| Feels quite tired | 95 (3.0) | 46 (3.1) | 49 (3.0) |
| Feels very tired | 43 (1.4) | 22 (1.5) | 21 (1.3) |
| Missing <sup>a</sup> | 100 (3.2) | 49 (3.3) | 51 (3.1) |
| **My child feels <u>annoyed</u> (today)** |  |  |  |
| Doesn't feel annoyed | 2412 (76.7) | 1115 (75.0) | 1297 (78.3) |
| Feels a little bit annoyed | 439 (14.0) | 229 (15.4) | 210 (12.7) |
| Feels a bit annoyed | 111 (3.5) | 53 (3.6) | 58 (3.5) |
| Feels quite annoyed | 41 (1.3) | 20 (1.3) | 21 (1.3) |
| Feels very annoyed | 24 (0.8) | 11 (0.7) | 13 (0.8) |
| Missing <sup>a</sup> | 116 (3.7) | 58 (3.9) | 58 (3.5) |
| **My child has problems with <u>Schoolwork/Homework</u> (today)** |  |  |  |
| No problems with schoolwork/homework | 2367 (75.3) | 1104 (74.3) | 1263 (76.2) |
| A few problems with schoolwork/homework | 472 (15.0) | 228 (15.3) | 244 (14.7) |
| Some problems with schoolwork/homework | 138 (4.4) | 68 (4.6) | 70 (4.2) |
| Many problems with schoolwork/homework | 45 (1.4) | 20 (1.3) | 25 (1.5) |
| Can't do schoolwork/homework | 20 (0.6) | 12 (0.8) | 8 (0.5) |
| Missing <sup>a</sup> | 100 (3.2) | 53 (3.6) | 47 (2.8) |
| **My child has problems with <u>Sleep</u> (last night)** |  |  |  |
| Had no problems sleeping | 2524 (80.3) | 1171 (78.8) | 1353 (81.7) |
| Had a few problems sleeping | 383 (12.2) | 188 (12.7) | 195 (11.8) |
| Had some problems sleeping | 106 (3.4) | 56 (3.8) | 50 (3.0) |
| Had many problems sleeping | 40 (1.3) | 22 (1.5) | 18 (1.1) |
| Couldn't sleep at all | 4 (0.1) | 3 (0.2) | 1 (0.1) |
| Missing <sup>a</sup> | 85 (2.7) | 45 (3.0) | 40 (2.4) |
| **My child has problems with <u>Daily routine</u> (today)** |  |  |  |
| No problems with daily routine | 2625 (83.5) | 1229 (82.7) | 1396 (84.2) |
| A few problems with daily routine | 330 (10.5) | 164 (11.0) | 166 (10.0) |
| Some problems with daily routine | 76 (2.4) | 36 (2.4) | 40 (2.4) |
| Many problems with daily routine | 23 (0.7) | 10 (0.7) | 13 (0.8) |
| Can't do daily routine | 5 (0.2) | 2 (0.1) | 3 (0.2) |
| Missing <sup>a</sup> | 84 (2.7) | 45 (3.0) | 39 (2.4) |
| **My child can join in <u>Activities</u> (today)** |  |  |  |
| Can join in with any activities | 2648 (84.3) | 1240 (83.4) | 1408 (85.0) |
| Can join in with most activities | 268 (8.5) | 122 (8.2) | 146 (8.8) |
| Can join in with some activities | 95 (3.0) | 54 (3.6) | 41 (2.5) |
| Can join in with a few activities | 33 (1.0) | 21 (1.4) | 12 (0.7) |
| Can join in with no activities | 14 (0.4) | 5 (0.3) | 9 (0.5) |
| Missing <sup>a</sup> | 85 (2.7) | 44 (3.0) | 41 (2.5) |
| **Health Related Quality of Life (Utility Score) <sup>b</sup>** |  |  |  |
| Non-missing | 2968 (94.4) | 1400 (94.2) | 1568 (94.6) |
| Score based on Child Health Utility 9D Questionnaire, mean +/- SD | 0.96 +/- 0.06 | 0.96 +/- 0.05 | 0.96 +/- 0.06 |
| Missing <sup>a</sup> | 175 (5.6) | 86 (5.8) | 89 (5.4) |

<sup>a</sup> Not reported or not known (as % of the returned questionnaires); <sup>b</sup> Calculated for respondents with all domains reported; SD, standard deviation

The overall QoL utility score based on CHU9D was similar between the two sites, 0.96 (SD=0.05) for London and 0.96 (SD=0.06) for Luton. Due to the questions not being answered, the QoL utility score could not be calculated for 86 (5.8%) questionnaire respondents in London and 89 (5.4%) respondents in Luton.

### Resources used and costs

The parent/carer responses to questions on their child’s school absences due to ill health and resulting childcare costs and spending on medication/equipment at baseline are summarised in Table 7. The proportion of respondents that had to pay for childcare due to their child’s respiratory ill health was 3.8% (N=56) in London and 3.1% (N=52) in Luton. Less than half of respondents declared they had to pay out-of-pocket for medication/equipment (N=615; 41.4% in London vs N=796; 48.0% in Luton). The mean parent-reported costs for childcare in the last 12 months were somewhat higher in London £188.30/child (SD £169.60) compared to Luton £164.30/child (SD £182.30), while the mean amount spent on medicines was higher in Luton £73.40 (SD £353.80) than in London £65.80 (SD £137.10).

**Table 7.** Proportion of parents/carers paying for childcare and medication/equipment in the last 12 months due to their child’s respiratory ill health and the mean costs per respondent, data are N (% of those who returned a questionnaire)

| Variable | All | London | Luton |
| --- | --- | --- | --- |
| **Paying for medical costs** |  |  |  |
| Medical: Did not pay | 1364 (43.4) | 660 (44.4) | 704 (42.5) |
| Medical: Paid privately | 1411 (44.9) | 615 (41.4) | 796 (48.0) |
| Medical: Missing <sup>a</sup> | 368 (11.7) | 211 (14.2) | 157 (9.5) |
| **Medical: Mean costs (last 12 months)** |  |  |  |
| Medical: Paid privately (positive amount declared) <sup>b</sup> | 730 (23.2) | 331 (22.3) | 399 (24.1) |
|  | 69.9 +/- 277.2 | 65.8 +/- 137.1 | 73.4 +/- 353.8 |
| Medical: Mean costs (£) +/- SD per child/last 12 months |  |  |  |
| **Paying for childcare** |  |  |  |
| Childcare: Did not pay | 2711 (86.3) | 1266 (85.2) | 1445 (87.2) |
| Childcare: Paid for childcare <sup>2</sup> | 108 (3.4) | 56 (3.8) | 52 (3.1) |
| Childcare: Missing <sup>a</sup> | 324 (10.3) | 164 (11.0) | 160 (9.7) |
| **Childcare: Mean costs (last 12 months)** |  |  |  |
| Childcare: Paid privately (positive amount declared) <sup>b</sup> | 46 (1.5) | 31 (2.1) | 15 (0.9) |
| Childcare: Mean costs (£) +/- SD per child/last 12 months | 180.5 +/- 172.2 | 188.3 +/- 169.6 | 164.3 +/- 182.3 |

<sup>a</sup> Not reported or not known (as % of the returned questionnaires); <sup>b</sup> For respondents who have reported a positive amount paid over 12 months; SD, standard deviation

Responses to questions on parents/carers taking time off work due to their child’s ill health are summarised in Table 8. More parents/carers reported taking time off due to their child’s ill health in London compared with Luton (23.9% vs 21.4% for any reason; 13.0% vs 8.9% for respiratory ill health).

**Table 8.**
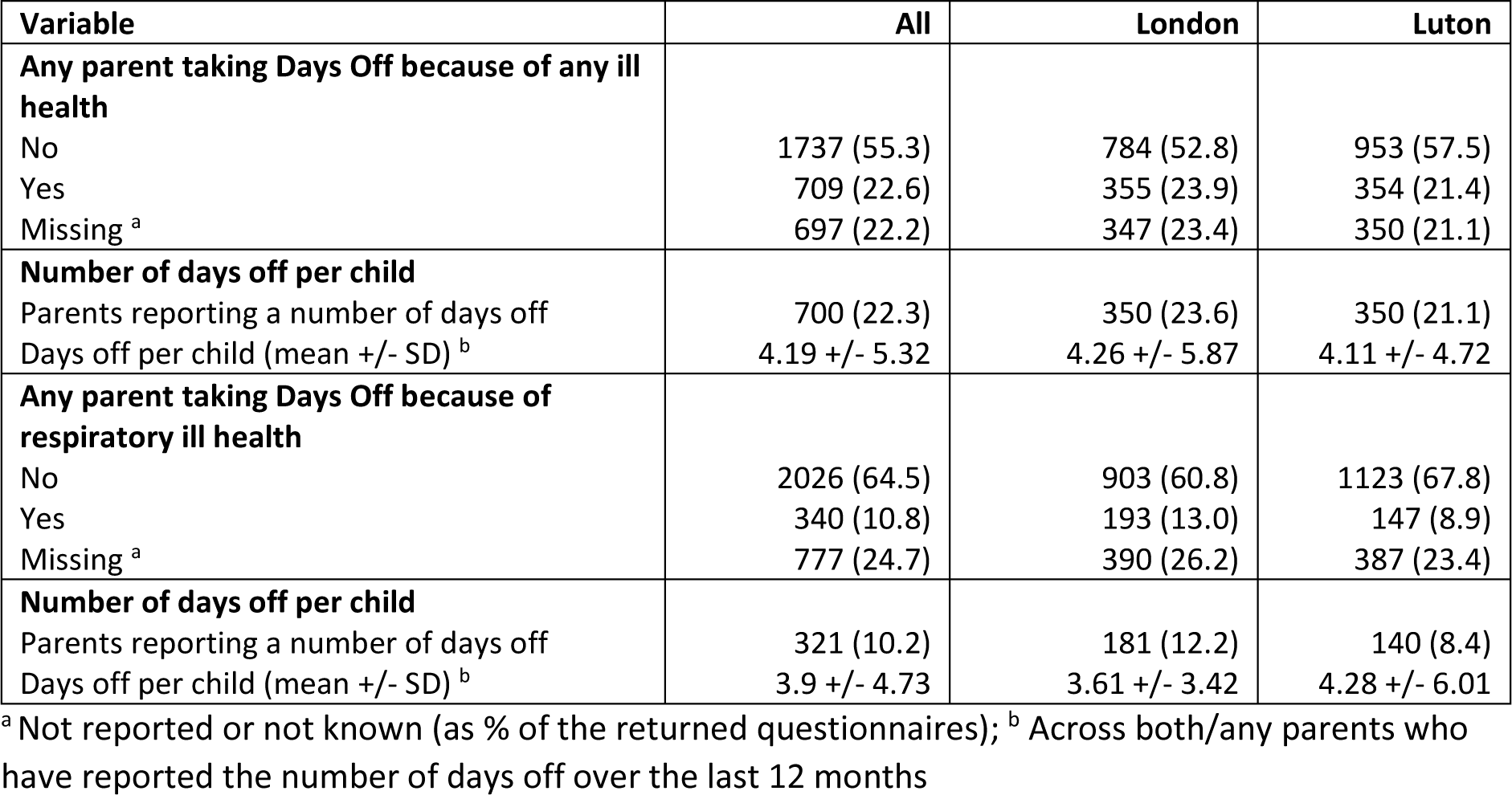
Proportions of parents/carers taking time off work due to their child’s ill health.

### Physical activity and GPS monitoring

Of 3414 children participating in CHILL, 2582 (76%) agreed to wear an accelerometer and 2176 (84%) provided valid accelerometery data (1258 participants from London and 918 participants from Luton). 835 (25%) participants also provided GPS data (439 participants from London and 396 participants from Luton). Baseline physical activity characteristics of the cohort are described in depth elsewhere (Scales et al, submitted).

### Air Quality

While Luton fulfilled many of the requirements for a suitable comparator site in this study, it is important to acknowledge that pollutant concentrations, specifically at road/kerbside locations were lower than in central London, and that monitoring within Luton and Dunstable was much less comprehensive at the beginning of the study. Mean annual concentrations (2017-2019) for NO_2_, particulate matter with a diameter of less than 2.5 micrograms (PM_2.5_) and less than 10 micrograms (PM_10_) at roadside and urban background sites within the ULEZ and in Luton are illustrated in Table 9. Roadside NO_2_ concentrations within the ULEZ zone were markedly higher than those in Luton, but this difference was strongly influenced by the kerbside sites in the ULEZ (particularly Marylebone Road 2018, 85 μg/m^3^; kerbsite sampling sites are within 1m of the kerb of a busy road, roadside sites are within 1-5m of a busy road). For 2018, no automated network NO_2_ data were available from a background site in Luton, but annual NO_2_ concentrations were available from diffusion tubes across a range of sites for 2018: roadside (35.1±5.1 μg/m^3^, mean±SD, n=20), suburban (27.9±2.3 μg/m^3^, n=9), and urban background (31.0±0.0 μg/m^3^, n=3). Therefore, background NO_2_ concentrations across the two sites appeared broadly similar. Higher roadside PM_2.5_ and PM_10_ concentrations were also observed with the ULEZ zone, compared with Luton, although these differences were less pronounced. PM_10_ concentrations were however similar at background sites at the two locations, Table 9. No background PM_2.5_ measurements were available in Luton in 2018.

**Table 9.** Annual mean (SD) pollutant concentrations from selected automatic air quality monitoring sites.

|  |  | NO<sub>2</sub> (mg/m<sup>3</sup>) |  |  |
| --- | --- | --- | --- | --- |
|  |  | 2017 | 2018 | 2019 |
| **London – ULEZ** | Roadside | 63.8±18.2 (n=4) | 60.6±16.7 (n=5) | 46.6±10.4 (n=7) |
|  | Urban background | 36.3±1.7 (n=4) | 34.5±3.7 (n=4) | 33.8±3.9 (n=4) |
| **Luton** | Roadside | 41.5±3.5 (n=2) | 40.0±4.2 (n=2) | 39.5±0.7 (n=2) |
|  | Urban background | NA | NA | NA |
|  |  | PM<sub>2.5</sub> (mg/m<sup>3</sup>) |  |  |
|  |  | 2017 | 2018 | 2019 |
| **London – ULEZ** | Roadside | 13.4±2.3 (n=2) | 13.1±4.1 (n=2) | 11.6±3.5 (n=2) |
|  | Urban background | 11.0±2.8 (n=2) | 10.5±0.7 (n=2) | 11.5±0.7 (n=2) |
| **Luton** | Roadside | 10.0 (n=1) | 10.0 (n=1) | 10.0 (n=1) |
|  | Urban background | NA | NA | NA |
|  |  | PM<sub>10</sub> (mg/m<sup>3</sup>) |  |  |
|  |  | 2017 | 2018 | 2019 |
| **London – ULEZ** | Roadside | 23.0±1.0 (n=3) | 25.2±2.7 (n=5) | 24.0±2.1 (n=5) |
|  | Urban background | 18.3±1.2 (n=3) | 18.0±1.7 (n=3) | 17.3±0.6 (n=3) |
| **Luton** | Roadside | 16.0 (n=1) | 16.0 (n=1) | 16.0 (n=1) |
|  | Urban background | 18.0 (n=1) | 17.0 (n=1) | 15.0 (n=1) |

**London roadside sites:** Westminster Duke Street (NO<sub>2</sub>, 2019); Southwark, Tower Bridge Road (NO<sub>2</sub>, 2017, 18, 19; PM<sub>10</sub>, 2017, 18, 19); Hackney, Old Street (NO<sub>2</sub>, 2017, 18, 19; PM<sub>2.5</sub>, 2017, 18, 19; PM<sub>10</sub>, 2017, 18, 19); Westminster, Duke Street (NO<sub>2</sub>, 2019); Westminster, Marylebone Road (NO<sub>2</sub>, 2017, 18, 19; PM<sub>2.5</sub>, 2017, 18, 19; PM<sub>10</sub>, 2017, 18, 19); Westminster, Cavendish Square (NO<sub>2</sub>, 2018, 19; PM<sub>10</sub>, 2018, 19); Westminster, Oxford Street (NO<sub>2</sub>, 2017, 18, 19; PM<sub>10</sub>, 2018, 19). **London urban background sites:** Westminster, Horseferry Road (NO<sub>2</sub>, 2017, 18, 19; PM<sub>2.5</sub>, 2017, 18, 19; PM<sub>10</sub>, 2017, 18, 19); Westminster, Covent Garden (NO<sub>2</sub>, 2017, 18, 19); Southway, Elephant and Castle (NO<sub>2</sub>, 2017, 18, 19; PM<sub>10</sub>, 2017, 18, 19); Camden, Bloomsbury square (NO<sub>2</sub>, 2017, 18, 19; PM<sub>2.5</sub>, 2017, 18, 19; PM<sub>10</sub>, 2017, 18, 19). **Luton/Dunstable roadside sites:** Luton A505 (NO<sub>2</sub>, 2017, 18, 19); Luton Dunstable Road East (NO<sub>2</sub>, 2017, 18, 19; PM<sub>2.5</sub>, 2017, 18, 19; PM<sub>10</sub>, 2017, 18, 19). **Luton/Dunstable background sites:** Luton Airport (PM<sub>10</sub>, 2017, 18, 19). **NA:** data not available. Data sources: (24), (25), (26), (27), (28).

For CHILL, individual pollutant exposures will be estimated based on high resolution monthly and annual models. Estimates will be derived from the CMAQ (Community Multiscale Air Quality Modeling System) urban model, which combines the ADMS (Advanced Dispersion Modelling System) roads model, WRF (Weather Research and Forecasting) meteorological model, and CMAQ regional scale models (29), to estimate exposures to various air pollutants, including NOx, NO_2_, O_3_, SO_2_, PM_2.5_, PM_10_, and PM components – primary and secondary inorganic and organic fractions. Exposure estimates will be produced at resolutions of up to 20m² in urban areas of the UK. Model surfaces for NO_2_, PM_10_ and PM_2.5_ for 2018 across the two study areas are illustrated in Fig. 6.

**Fig. 6.**
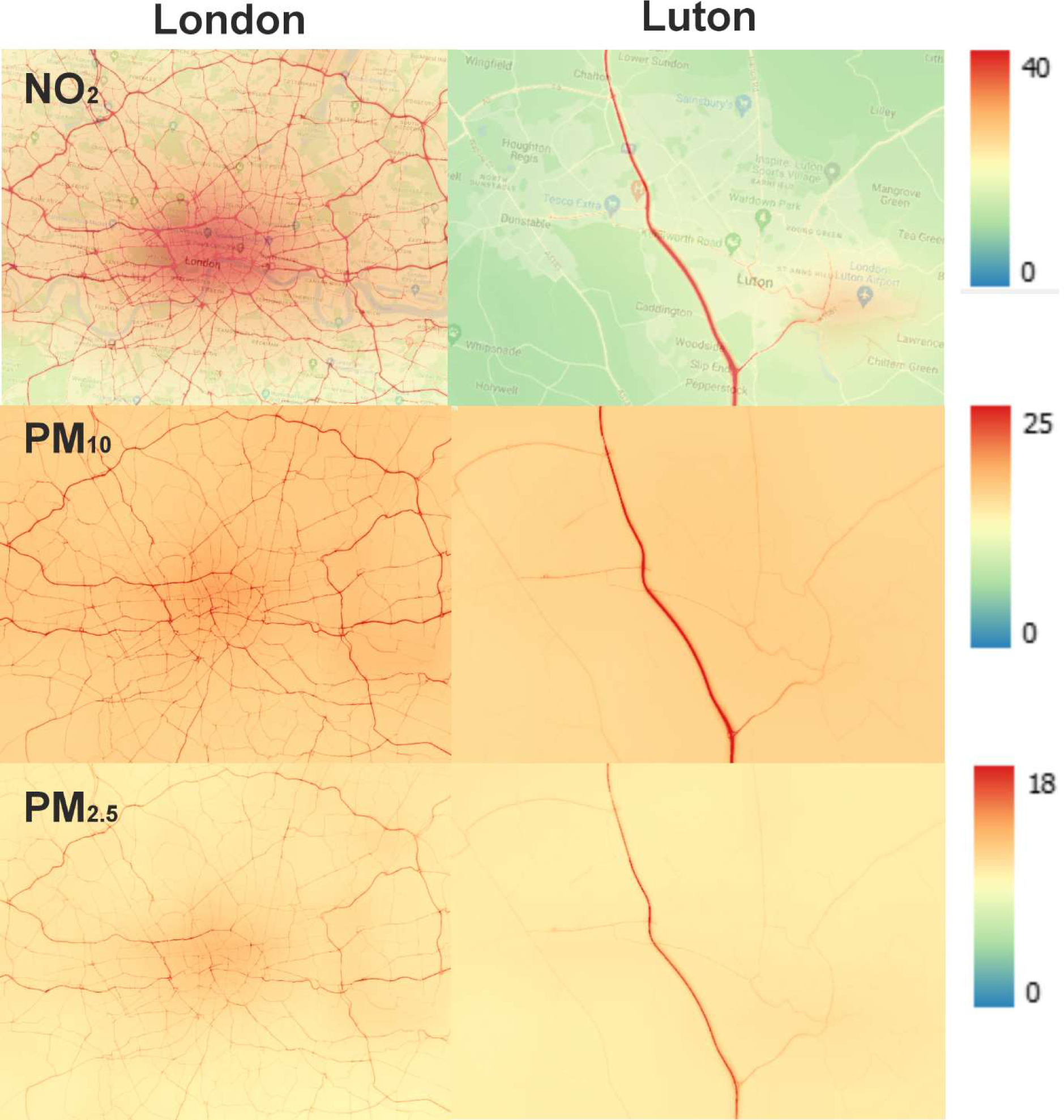
Modelled annual pollutant concentrations for nitrogen dioxide (NO_2_) and particulate matter with a diameter of less than 10 micrograms (PM_10_) and less than 2.5 micrograms (PM_2.5_) across London and Luton study sites for 2018. All concentrations expressed as μg/m^3^

## Discussion

### Summary

We successfully established a cohort of 3414 primary school children drawn from 84 primary schools in central London and Luton with the aim of evaluating the health impacts of AQ improvements following the implementation of the ULEZ. The study is a natural experiment evaluation, comparing effects over five years in two populations: children living in London exposed to the effects of the ULEZ, and children living in Luton, which has no low emission zone.

The demography of participating children is broadly similar across the two sites, with both including children from a range of ethnic groups, of similar mean age, and many living in areas of socioeconomic disadvantage. The two populations are closely matched in terms of lung function (the primary outcome is lung function growth as measured by FEV_1_), as well as a wide range of anthropomorphic, behavioural, and quality of life variables. Lung function data were of high quality, with a high proportion of children achieving successful spirometric assessments, despite their young age. Reported prevalences of asthma, eczema and rhinitis are similar to those seen in our previous work in children of similar ages in London (30). Travel behaviours differed slightly across the two sites, with more children in London using active travel, probably reflecting shorter journeys to school than those of the children in Luton. Air quality at background monitoring sites were broadly similar in the two study areas, however NO_2,_ and to a lesser extent PM_2.5_ and PM_10_, concentrations were notably higher at London roadside locations. These differences, particular those for NO_2_, are likely driven by pollution “hot-spots” such as Marylebone Road. The study population health assessments and AQ measurements were made in a timely way, immediately before the implementation of the ULEZ in April 2019, giving valuable pre-intervention data, prior to further assessments over the course of the study. We exceeded the recruitment target of 3200 children, which gives the study a high chance of maintaining the power to answer the primary study question of the impacts of AQ improvements on lung growth, as well as carrying out pre-specified sub-group analyses, including effects on health inequalities.

### Strengths and weaknesses

We have chosen a parallel prospective cohort design. To our knowledge, this approach is unique in the field of evaluating a city-wide air pollution intervention. Merits of this approach, using a parallel geographically distinct comparator population, include the ability to more accurately account for secular trends in AQ arising from national level policies, for example, subsidies to incentivise a switch to electric vehicles, and to account, at least partially, for unexpected population-wide disruption, including the COVID-19 pandemic.

There is a growing interest in the challenge of evaluating population health interventions, where randomised designs are often impossible. Ogilvie and colleagues argue that natural experiment designs are not inferior to randomised trials and are ‘the only way to generate meaningful evidence to address critical questions about investing in population health interventions’ (31).

Our study collects outcomes from multiple sources, including direct physiological assessment (e.g., lung function), behavioural (accelerometry, GPS), parental and child self-report (e.g., symptoms, travel behaviours, quality of life), and health record data (respiratory infection and new diagnoses). This combination of data allows both triangulation across sources, and elucidation of possible mechanistic pathways that influence health.

High quality longitudinal AQ data is a particular strength of the study. Additional monitoring was set up at the start of the CHILL Study for the Luton site. Both sites give AQ data for criterion pollutants.

The cohort provides a robust platform on which to add sub-studies, including qualitative analyses to explore behavioural influences, choices, and mechanisms, gathering additional outcomes to address additional hypotheses, measuring additional confounding variables, as well as potentially capturing the impact of further extensions of the ULEZ.

A potential weakness of the study is attrition of the cohort over time, although a 20% annual loss is factored into power estimates, and the study team have made significant efforts to encourage ongoing participation, both by schools and individual participants. Such efforts have included: maintaining contact with the schools between annual health assessments; providing new educational outreach workshops for each year of the study, focusing on different aspects of the effects of air pollution on health; providing a small monetary incentive for completion of the parent/carer questionnaire; giving each participant a CHILL-branded pen and badge in a different colour for each year of the study; and contacting parents to trace children moving up to secondary school. It is possible that implementation of the ULEZ may have affected the housing market and/or personal finances prompting families to move out of London, and there may also have been some impact of people moving out of cities and/or returning to their home countries during the COVID pandemic.

In summary, the CHILL cohort comprises a large sample of ethnically diverse children living in areas of London and Luton with high levels of TRAP, well matched for high quality pre-intervention lung function data and demographic variables, with parallel high quality measurement of AQ. Using a prospective parallel cohort natural experiment design, the cohort provides an outstanding platform for the evaluation of the impact of London’s ULEZ on children’s lung function growth and respiratory health.

## Data Availability

All data produced in the present study are available upon reasonable request to the authors

## STATEMENTS AND DECLARATIONS

## Funding

This work was supported by the National Institute of Health Research Public Health Research (grant number 16/139/09) with additional funding by NIHR CLAHRC North Thames, Barts Charity, and NIHR ARC North Thames. EvS is funded by the Medical Research Council [MC UU 00006/5]. BM acknowledges support by the National Institute for Health Research Barts Biomedical Research Centre (NIHR203330). CJG is supported by the NIHR ARC North Thames. The views expressed in this publication are those of the authors and not necessarily those of the NIHR or the Department of Health and Social Care. Funders played no role in the design of the study and collection, analysis, and interpretation of data and in writing the manuscript.

## Competing interests

The authors have no relevant financial or non-financial interests to disclose.

## Authors’ contributions

The study was designed by Chris J. Griffiths, Ian S. Mudway, Aziz Sheikh, Frank Kelly, Gurch Randhawa, Borislava Mihaylova, Esther van Sluijs, Jonathan Grigg, W. James Gauderman, Helen E. Wood, James Scales, Rosamund E. Dove, Louise Cross, Ivelina Tsocheva, Harpal Kalsi and Bill Day. The first draft of the manuscript was written by Helen E. Wood, with the section on air quality being written by Ian S. Mudway (data provided by Sean Beevers), and the sections on quality of life and resource use being written by Florian Tomini, Veronica Tofolutti and Borislava Mihaylova. All authors commented on previous versions of the manuscript. All authors read and approved the final manuscript.

## Compliance with ethical standards

Ethics approval for research on human participants has been given by Queen Mary University of London Research Ethics Committee (ref 2018/08), and NHS West of Scotland Research Ethics Committee 4 (reference 22/WS/0065). All methods were carried out in accordance with relevant guidelines and regulations. Informed consent forms were completed by caregivers and children at home. Written informed consent was obtained from the participants’ parent/carer. Participants were also asked for their assent.

## Data availability

The datasets used and/or analysed during the current study are available from the corresponding author upon reasonable request.

## Acknowledgements

We are grateful to all participating schools, parents and children for their enthusiasm and support. The views expressed in this paper are those of the authors and should not be taken to reflect the official position of the funder. This report contains independent research supported by the National Institute for Health and Care Research ARC North Thames. The views expressed in this publication are those of the author(s) and not necessarily those of the National Institute for Health and Care Research or the Department of Health and Social Care. IM, FJK and SB were supported by the NIHR Health Protection Research Units in Environmental Exposures and Health, and Chemical and Radiation Threats and Hazards, a partnership between the UK Health Security Agency and Imperial College London. The views expressed are those of the authors and not necessarily those of the NIHR, UKHSA, or the Department of Health and Social Care.

For the purpose of Open Access, the author has applied a Creative Commons Attribution (CC BY) licence to any Author Accepted Manuscript version arising.

## Notes

### Competing Interest Statement

The authors have declared no competing interest.

### Clinical Protocols

https://pubmed.ncbi.nlm.nih.gov/37925402/

### Funding Statement

This study was supported by the National Institute of Health Research Public Health Research (grant number 16/139/09) with additional funding by NIHR CLAHRC North Thames, Barts Charity, and NIHR ARC North Thames. EvS is funded by the Medical Research Council [MC UU 00006/5]. BM acknowledges support by the National Institute for Health Research Barts Biomedical Research Centre (NIHR203330). CJG is supported by the NIHR ARC North Thames. The views expressed in this publication are those of the authors and not necessarily those of the NIHR or the Department of Health and Social Care. Funders played no role in the design of the study and collection, analysis, and interpretation of data and in writing the manuscript.

### Author Declarations

Queen Mary University of London Research Ethics Committee (ref 2018/08) and NHS West of Scotland Research Ethics Committee 4 (reference 22/WS/0065) gave ethical approval for this work.

## References

1. Holgate ST. ’Every breath we take: the lifelong impact of air pollution’ - a call for action. Clin Med (Lond). 2017;17(1):8–12. doi:10.7861/clinmedicine.17-1-8

2. WHO. 7 million premature deaths annually linked to air pollution. https://www.who.int/news/item/25-03-2014-7-million-premature-deaths-annually-linked-to-air-pollution2014.

3. Lelieveld J, Klingmüller K, Pozzer A, et al. Cardiovascular disease burden from ambient air pollution in Europe reassessed using novel hazard ratio functions. Eur Heart J. 2019;40(20):1590–6. doi:10.1093/eurheartj/ehz135

4. Barnes JH, Chatterton TJ, Longhurst JW. Emissions vs exposure: Increasing injustice from road traffic-related air pollution in the United Kingdom. Transportation research part D: transport and environment. 2019;73:56–66. doi:10.1016/j.trd.2019.05.012

5. Fecht D, Fischer P, Fortunato L, et al. Associations between air pollution and socioeconomic characteristics, ethnicity and age profile of neighbourhoods in England and the Netherlands. Environ Pollut. 2015;198:201–10. doi:10.1016/j.envpol.2014.12.014

6. Landrigan PJ, Fuller R, Acosta NJR, et al. The Lancet Commission on pollution and health. Lancet. 2018;391(10119):462–512. doi:10.1016/s0140-6736(17)32345-0

7. Suk WA, Ahanchian H, Asante KA, et al. Environmental Pollution: An Under-recognized Threat to Children’s Health, Especially in Low- and Middle-Income Countries. Environ Health Perspect. 2016;124(3):A41–5. doi:10.1289/ehp.1510517

8. Liu NM, Grigg J. Diesel, children and respiratory disease. BMJ Paediatr Open. 2018;2(1):e000210. doi:10.1136/bmjpo-2017-000210

9. Wang J, Cao H, Sun D, et al. Associations between ambient air pollution and mortality from all causes, pneumonia, and congenital heart diseases among children aged under 5 years in Beijing, China: A population-based time series study. Environ Res. 2019;176:108531. doi:10.1016/j.envres.2019.108531

10. HEI. COMMUNICATION 11 EXECUTIVE SUMMARY Assessing Health Impact of Air Quality Regulations: Concepts and Methods for Accountability Research. https://www.healtheffects.org/system/files/Comm11ExecSumm.pdf2003.

11. Tsocheva I, Scales J, Dove R, et al. Investigating the impact of London’s ultra low emission zone on children’s health: children’s health in London and Luton (CHILL) protocol for a prospective parallel cohort study. BMC Pediatr. 2023;23(1):556. doi:10.1186/s12887-023-04384-5

12. ClinicalTrials.gov. Children’s Health in London and Luton (CHILL). 2021. https://clinicaltrials.gov/study/NCT04695093.

13. Colligan G, Tsocheva I, Scales J, et al. Investigating the impact of London’s Ultra Low Emission Zone on children’s health: Children’s Health in London and Luton (CHILL): Protocol for a prospective parallel cohort study. medRxiv. 2021:2021.02.04.21251049. doi:10.1101/2021.02.04.21251049

14. Asher MI, Keil U, Anderson HR, et al. International Study of Asthma and Allergies in Childhood (ISAAC): rationale and methods. Eur Respir J. 1995;8(3):483–91. doi:10.1183/09031936.95.08030483

15. Stevens K. Valuation of the Child Health Utility 9D Index. Pharmacoeconomics. 2012;30(8):729–47. doi:10.2165/11599120-000000000-00000

16. Ministry of Housing CLG. English indices of deprivation 2019. 2019. https://www.gov.uk/government/statistics/english-indices-of-deprivation-2019.

17. Graham BL, Steenbruggen I, Miller MR, et al. Standardization of Spirometry 2019 Update. An Official American Thoracic Society and European Respiratory Society Technical Statement. Am J Respir Crit Care Med. 2019;200(8):e70-e88. doi:10.1164/rccm.201908-1590ST

18. Miller MR, Hankinson J, Brusasco V, et al. Standardisation of spirometry. Eur Respir J. 2005;26(2):319-38. doi:10.1183/09031936.05.00034805

19. Sylvester KP, Clayton N, Cliff I, et al. ARTP statement on pulmonary function testing 2020. BMJ Open Respir Res. 2020;7(1). doi:10.1136/bmjresp-2020-000575

20. Luton Council. 2021 Census ethnicity language nationality and religion. 2024. https://m.luton.gov.uk/Page/Show/Community_and_living/Luton%20observatory%20census%20statistics%20and%20mapping/population/Pages/2021-Census-ethnicity-language-nationality-and-religion.aspx#:∼:text=The%20white%20ethnic%20group%20makes,of%20the%20population%20of%20Luton. Accessed 19/02/2024.

21. Luton Borough Council. Luton Borough Council Air Quality Action Plan.2018.

22. Luton Borough Council. 2021 Air Quality Annual Status Report (ASR).2021.

23. Quanjer PH, Stanojevic S, Cole TJ, et al. Multi-ethnic reference values for spirometry for the 3-95-yr age range: the global lung function 2012 equations. Eur Respir J. 2012;40(6):1324–43. doi:10.1183/09031936.00080312

24. Luton Borough Council. 2020 Air Quality Annual Status Report (ASR).2020.

25. London Borough of Camden. London Borough of Camden Air Quality Annual Status Report for 2020. 2020 July 2021.

26. London Borough of Hackney. London Borough of Hackney Air Quality Annual Status Report for 2021.2021 May 2022.

27. Southwark Council. Southwark Council Air Quality Annual Status Report for 20212021 May 2022.

28. Westminster City Council. Westminster City Council Air Quality Annual Status Report for 2021.2021.

29. Beevers SD, Kitwiroon N, Williams ML, Kelly FJ, Ross Anderson H, Carslaw DC. Air pollution dispersion models for human exposure predictions in London. J Expo Sci Environ Epidemiol. 2013;23(6):647–53. doi:10.1038/jes.2013.6

30. Wood HE, Marlin N, Mudway IS, et al. Effects of Air Pollution and the Introduction of the London Low Emission Zone on the Prevalence of Respiratory and Allergic Symptoms in Schoolchildren in East London: A Sequential Cross-Sectional Study. PLoS One. 2015;10(8):e0109121. doi:10.1371/journal.pone.0109121

31. Ogilvie D, Adams J, Bauman A, et al. Using natural experimental studies to guide public health action: turning the evidence-based medicine paradigm on its head. Journal of Epidemiology and Community Health. 2020;74(2):203–8. doi:10.1136/jech-2019-213085

